# Cost-effectiveness of recombinant influenza vaccine compared with standard-dose influenza vaccine among adults 50 years of age and older in Hong Kong

**DOI:** 10.64898/2026.03.07.26347845

**Authors:** Shuyi Zhong, Irene O. L. Wong, Peng Wu, Benjamin J. Cowling

## Abstract

**Background:** Older adults face a disproportionately high risk of severe influenza, yet the standard inactivated vaccine (IIV) offers suboptimal protection in this population. This study evaluates the cost-effectiveness of replacing IIV with the recombinant influenza vaccine (RIV) for adults aged ≥50, ≥65, and ≥80 years in Hong Kong.

**Methods:** A decision tree model was used to compare RIV with IIV for adults aged ≥50, ≥65, and ≥80 years in Hong Kong, from a societal perspective. Costs, quality-adjusted life-years (QALYs), and incremental cost-effectiveness ratios (ICERs) were evaluated with the impact of parameter uncertainty on the results assessed via deterministic and probabilistic sensitivity analyses.

**Results:** For adults ≥50 years, RIV increased total costs by USD5.1 (HKD39.8) per person and gained 0.00012 QALYs (ICER: USD40,659 [HKD317,140] per QALY) compared to IIV. Among adults ≥65 years, RIV cost USD6.0 (HKD46.8) more and gained 0.00021 QALYs (ICER: USD29,077 [HKD226,801] per QALY). For adults ≥80 years, RIV cost USD3.2 (HKD25.0) more and gained 0.00015 QALYs (ICER: USD21,092 [HKD164,518] per QALY). ICERs were less than willingness-to-pay thresholds of one to three times Hong Kong’s gross domestic product per capita.

**Conclusions:** RIV is cost-effective compared with IIV for adults ≥50, ≥65, and ≥80 years in Hong Kong, with the lowest ICER observed in individuals ≥80 years.

## Introduction

Older adults carry a disproportionately high risk of severe influenza, making them a priority for vaccination. In Hong Kong, persons aged 65 years and above accounted for most excess deaths related to seasonal influenza [1,2]. However, the inactivated influenza vaccine (IIV), the formulation most frequently administered, produces a weaker immune response and offers lower efficacy in older individuals than in younger healthier groups [3,4]. To close this protection gap, several enhanced influenza vaccines have been developed [5]. In 2022, the U.S. Advisory Committee on Immunization Practices recommended that adults 65 years and older preferentially receive one of these enhanced options: an adjuvanted vaccine, a high-dose inactivated vaccine, or a recombinant influenza vaccine (RIV) [6]. In Hong Kong, community-dwelling adults aged ≥50 years may receive either IIV or RIV, while RIV is preferentially recommended by the Hong Kong Center for Health Protection for older adults residing in long-term care facilities [7].

The recommendations that favor RIV over IIV among older adults are based on clinical data demonstrating its superior clinical protection in this population. Unlike egg-based IIV, RIV is produced in insect cells, yielding recombinant hemagglutinin without any extraneous egg proteins. This egg-free technology prevents the mutations that can arise during egg propagation and reduce vaccine effectiveness [8,9]. According to a randomized, double-blind, multicenter trial among adults aged 50 and older, RIV reduced the risk of laboratory-confirmed influenza illness by 31% compared with IIV [10]. In addition, RIV was more effective than IIV against laboratory-confirmed influenza hospitalization [11].

Beyond clinical protection, economic considerations are critical for informing vaccine policy. Although multiple studies have demonstrated that seasonal influenza vaccine programs are efficient across different countries and locations [12–15], no comparison has been made to evaluate the cost-effectiveness of RIV versus IIV among older adults. Therefore, this study aims to conduct a comparative cost-effectiveness analysis to estimate the incremental costs, quality-adjusted life-years (QALYs), and budgetary impact of adopting RIV in place of IIV among adults above various age thresholds (≥50, ≥65, and ≥80 years) in Hong Kong.

## Methods

We developed a decision tree model to estimate the cost-effectiveness and expected public health outcomes (self-care, outpatient care, hospitalization without intensive care unit (ICU) admission, hospitalization with ICU admission, and hospitalization followed by death) prevented by influenza vaccination with RIV compared with standard-dose IIV in various age groups including adults ≥50 years, ≥65 years, and ≥80 years. Costs were expressed in both USD and HKD, noting that 7.8 HKD is approximately 1 USD. The analyses were performed in both base case and sensitivity analyses. Our study received ethical approval from the Institutional Review Board of the University of Hong Kong.

### Model Structure and data sources

We developed a decision tree model to evaluate the cost-effectiveness of vaccination with RIV compared with IIV across different age strata. As shown in Figure 1, the model begins with two decision nodes representing the alternative vaccination strategies applied to each age stratum, followed by three chance nodes corresponding to: (1) the occurrence of influenza, (2) risk group classification, and (3) subsequent clinical outcomes following influenza virus infection. This model assumes no difference in the risk of acquiring influenza between high-risk and normal-risk individuals, but allows for differences in illness severity after infection [16]. The probabilities at each chance node were derived from data collected in Hong Kong or from the published literature. Base-case values for each parameter were set to the corresponding mean or median estimates. Where Hong Kong–specific data were unavailable, we used estimates from comparable East Asian settings or applied conservative assumptions. Parameters used in the model are summarized in Table 1.

**Figure 1.**
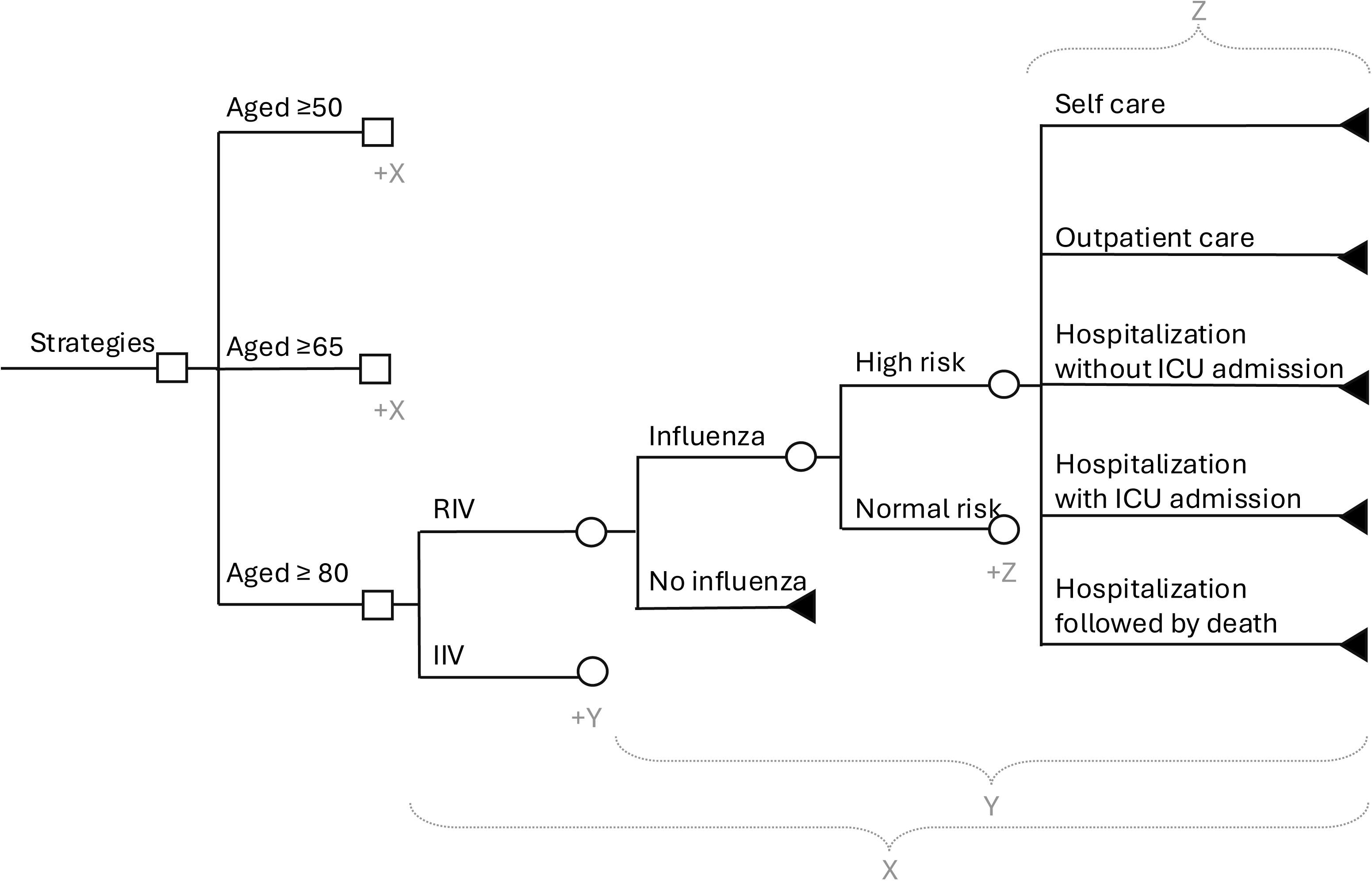
Decision tree model for evaluating the cost-effectiveness of influenza strategies among adults aged ≥50, ≥65, and ≥80 years in Hong Kong. The squares represent the decision nodes and circles represent chance nodes indicating probabilistic events, such as occurrence of influenza, risk group categorization, and subsequent clinical outcomes. Triangles represent terminal nodes, which correspond to final health outcomes. We repeated analyses in various age groups. Abbreviations: IIV, inactivated influenza vaccine; RIV, recombinant influenza vaccine; ICU, intensive care unit.

**Table 1.**
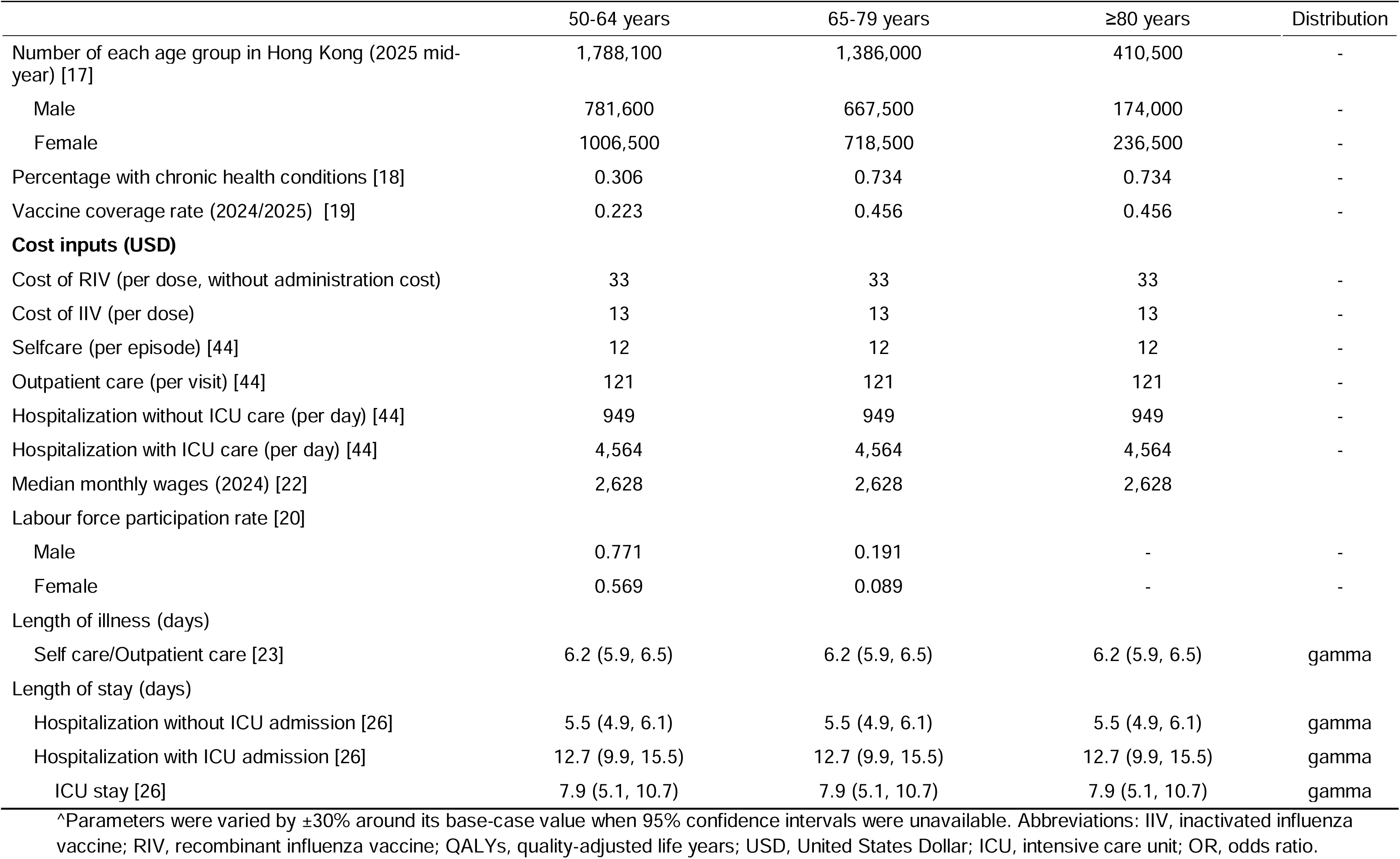
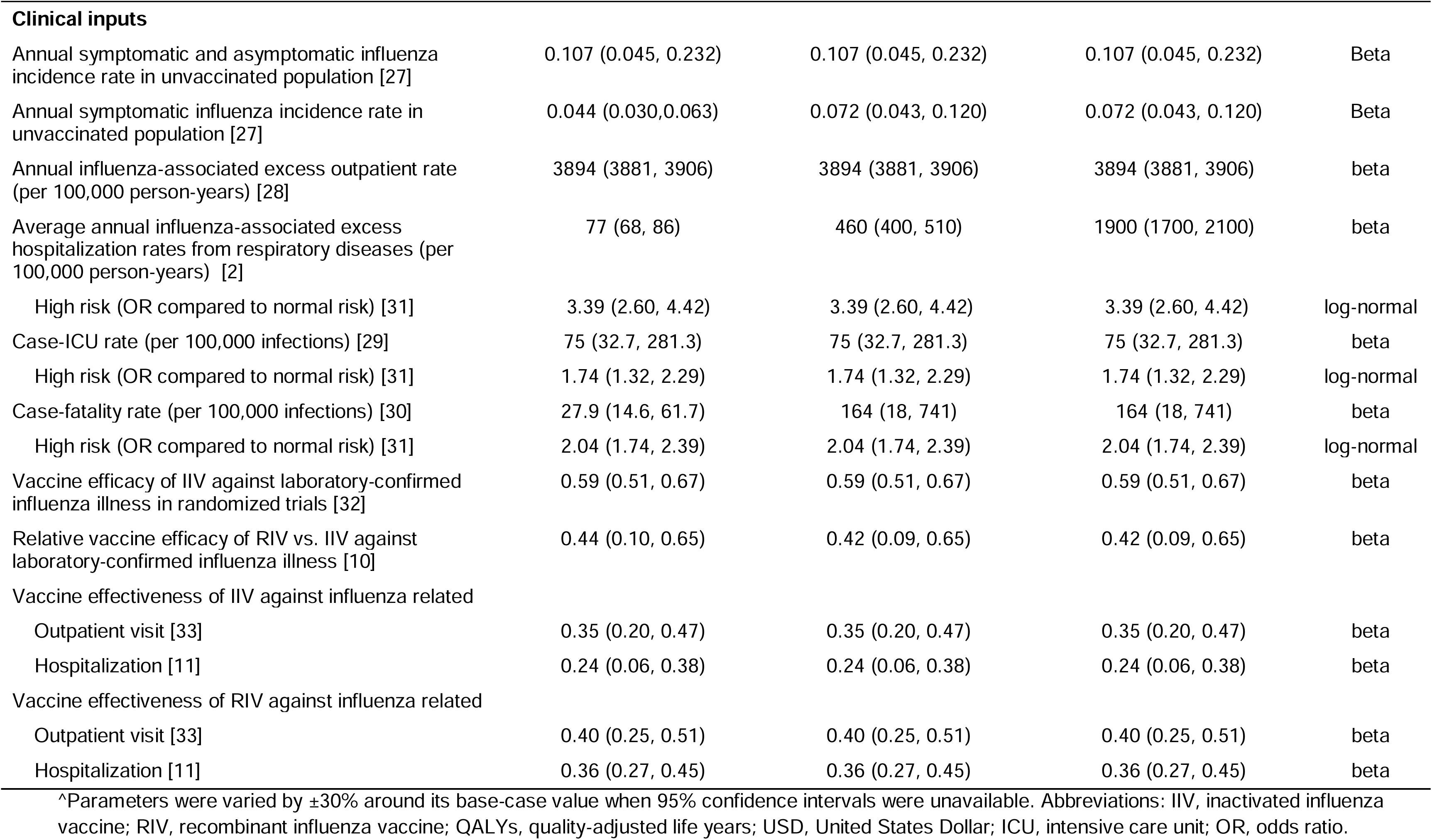
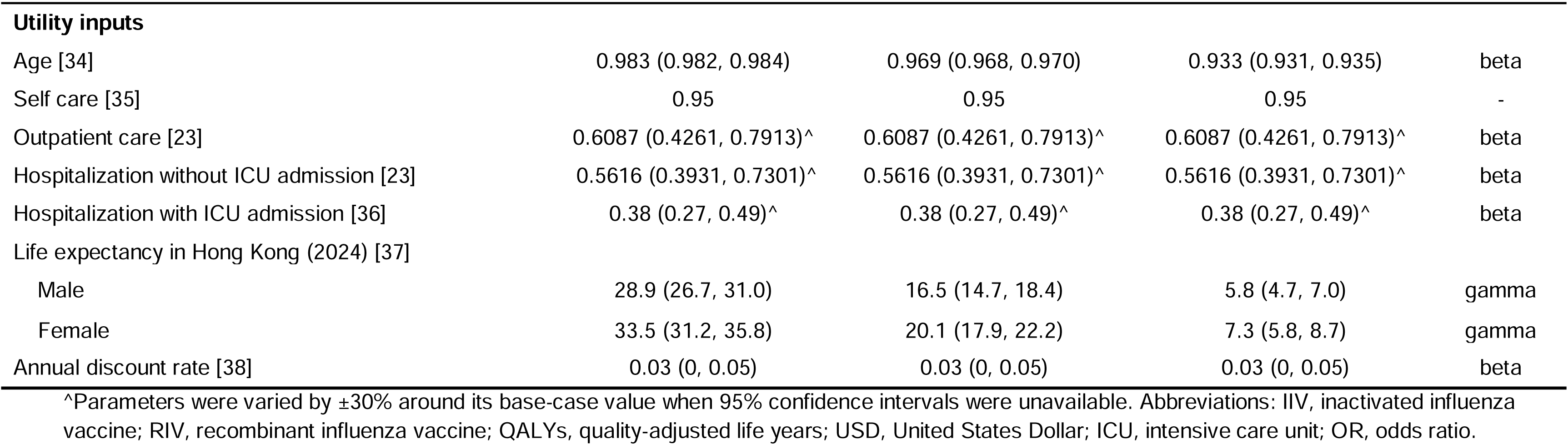
Parameters for calculation of costs and QALYs with RIV versus IIV.

The population figures for each age- and sex-specific group were obtained from the Census and Statistics Department [17]. High-risk populations for influenza were identified based on the presence of one or more chronic health conditions, as reported by the Health Bureau of the Hong Kong Government [18]. Vaccine coverage data were sourced from the Hong Kong Centre for Health Protection [19].

### Cost inputs

This analysis was conducted from a societal perspective, incorporating both direct and indirect costs. Direct costs included vaccination program expenses and medical costs for influenza-related illness. Indirect costs, including productivity losses due to influenza-related hospitalizations and deaths, were considered for adults under 80 years of age based on age-specific labour force participation rates [20]. Vaccine prices for RIV and IIV were provided by Sanofi. Administration fees were not considered because both comparison groups received vaccination.

Costs for self-medication, outpatient visits and hospitalizations (with and without ICU admission) were obtained from the Hospital Authority for patients not eligible for government subsidies, to reflect the full costs borne by both individuals and society [21]. The costs associated with death were assumed to be equivalent to those for hospitalization with ICU admission.

The median monthly wage was sourced from the Census and Statistics Department of Hong Kong [22]. The duration of illness was obtained from a retrospective telephone survey of laboratory-confirmed influenza cases from the 2013 National Influenza-like Illness Surveillance Network in China [23], and was assumed to be similar across self care and outpatient care settings [24,25]. Length of stay for hospitalization with and without hospitalizations with ICU admission was derived from a retrospective study of laboratory-confirmed influenza patients admitted to a tertiary hospital during 2014-2015 season in Hong Kong [26].

### Clinical inputs

The laboratory-confirmed symptomatic and asymptomatic influenza incidence rates among unvaccinated individuals were retrieved from a meta analysis that estimated attack rates of laboratory-confirmed seasonal influenza virus infections in the placebo groups of randomized controlled trials [27]. Since specific data on annual influenza-associated excess outpatient rates for the Hong Kong population were rarely reported, we used estimates from Japan as a proxy [28]. Average annual influenza-associated excess hospitalization rates from respiratory diseases were extracted from a study assessing disease burden of influenza in Hong Kong [2].

Case-ICU rate and case-fatality rate were derived from two surveillance studies conducted in Hong Kong in 2009 [29,30]. For the high-risk population, we assumed that incidence and outpatient rates were similar to those in the normal-risk population, while the annual influenza-associated excess hospitalization, case-ICU rate and case-fatality rate were calculated using the odds ratio (OR) for high-risk versus normal-risk groups obtained from a meta-analysis [31].

Vaccine effectiveness estimates for laboratory-confirmed influenza illness were unavailable, as observational studies typically report effectiveness against medically attended cases, categorized as outpatient care in this analysis. We therefore used vaccine efficacy against laboratory-confirmed influenza illness as a proxy for vaccine effectiveness. Because no data were identified on RIV efficacy versus placebo for laboratory-confirmed influenza illness, we inferred RIV efficacy using IIV efficacy from a randomized controlled trial in adults aged ≥50 years [32] combined with relative vaccine efficacy of RIV versus IIV from a meta-analysis [10]. Vaccine effectiveness against outpatient visit and hospitalization for IIV and RIV was extracted from test-negative design studies [11,33].

### Utility inputs

The QALYs gained by RIV versus IIV were calculated using the number of reduced outcomes of influenza illness, the utility value and duration of time-spent in each outcome. Age-specific health utility scores were sourced from the 2013 National Health Services Survey in China, representing the Chinese population [34]. Utility scores for self-care, outpatient care, and hospitalization (with and without ICU) were obtained from relevant literature [23,35,36]. Life expectancy in Hong Kong (2024) for different age groups was obtained from the Census and Statistics Department in Hong Kong [37]. To estimate QALYs gained from preventing non-fatal clinical outcomes, the number of events was multiplied by the duration of each outcome (in days) and the corresponding utility score dividing by 365 days. Future life years and productivity losses from deaths were discounted to present value using a 3% annual rate [38]. Discounting was applied to all costs and QALYs occurring beyond the first year, as the decision tree models a single influenza season but captures long-term mortality benefits.

### Cost-effectiveness analysis

The cost-effectiveness of RIV compared with IIV was determined by incremental cost-effectiveness ratios (ICER), which was calculated as follows:

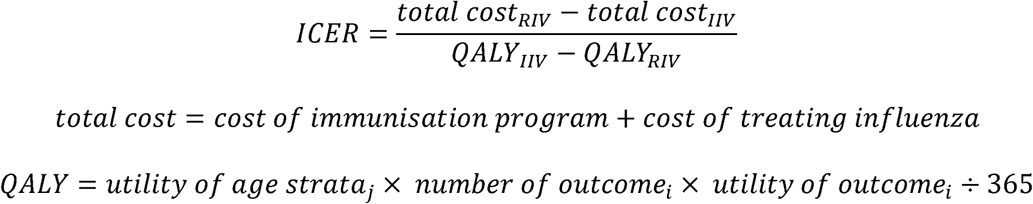

As recommended by the World Health Organization, interventions with an ICER below three times the gross domestic product (GDP) per capita are considered cost-effective [39]. Using the 2024 GDP per capita for Hong Kong SAR, China (USD54,101 [HKD421,990]), we therefore specified willingness-to-pay (WTP) thresholds at one to three times this value [40].

### Sensitivity analyses

Deterministic and probabilistic sensitivity analyses were conducted to assess the impact of parameter uncertainty on model outcomes in each age group (≥50, ≥65, and ≥80 years). In one-way sensitivity analyses, each parameter was varied individually across its 95% confidence interval; when this was unavailable, a ±30% range around the base-case value was applied. Sensitivity analysis results were presented as a tornado diagram. For the probabilistic sensitivity analysis, a Monte Carlo simulation was performed in which all parameters were varied simultaneously. Costs and QALYs were recalculated 1,000 times by randomly sampling each input from its assigned probability distribution (Table 1), and the resulting simulations were used to generate a cost-effectiveness plane. A cost-effectiveness acceptability curve was also constructed to show the probability that RIV, compared with IIV, is cost-effective across a range of WPT thresholds. All analyses were conducted in R version 4.3.1 (R Foundation for Statistical Computing, Vienna, Austria).

## Results

### Base case analysis

The impact of replacing IIV with RIV at different age thresholds on clinical outcomes, QALYs, and costs is summarized in Table 2. Among adults aged ≥50 years, we estimated that RIV would prevent an additional 13,322 influenza cases, 2,372 outpatient visits, 813 hospitalizations without ICU admission, 17 hospitalizations with ICU admission, and 17 deaths over 1 year, compared with IIV. Per person, RIV yielded gains of 0.00007 QALYs from fewer nonfatal episodes and 0.00006 QALYs from deaths averted. RIV increased vaccination costs by USD7.0 (HKD54.6) per person but reduced productivity losses and medical costs by USD0.2 (HKD1.56) and USD1.7 (HKD13.3) per person, respectively. Overall, total costs were USD5.1 (HKD39.8) higher per person with RIV than with IIV, for a net gain of 0.00012 QALYs per person.

**Table 2.** Disease outcomes and costs of RIV versus IIV in base-case analysis.

| Outcomes | Aged ≥50 years |  |  | Aged ≥65 years |  |  | Aged ≥80 years |  |  |
| --- | --- | --- | --- | --- | --- | --- | --- | --- | --- |
|  | IIV | RIV | Difference* | IIV | RIV | Difference* | IIV | RIV | Difference* |
| Total number of clinical outcomes |  |  |  |  |  |  |  |  |  |
| Self care | 31,376 | 18,054 | 13,322 | 24,183 | 14,026 | 10,157 | 5,526 | 3,205 | 2,321 |
| Outpatient care | 30,828 | 28,456 | 2,372 | 20,735 | 19,140 | 1,595 | 4,738 | 4,373 | 365 |
| Hospitalizations without ICU admission | 5,146 | 4,333 | 813 | 4,913 | 4,137 | 776 | 2,703 | 2,276 | 427 |
| Hospitalizations with ICU admission | 40 | 23 | 17 | 27 | 16 | 11 | 6 | 4 | 2 |
| Hospitalization followed by deaths | 41 | 24 | 17 | 40 | 23 | 17 | 9 | 5 | 4 |
| Health effects (per person) |  |  |  |  |  |  |  |  |  |
| QALYs lost due to non-fatal episodes | 0.00023 | 0.00017 | 0.00007 | 0.00035 | 0.00025 | 0.00010 | 0.00037 | 0.00026 | 0.00010 |
| QALYs lost due to deaths | 0.00014 | 0.00008 | 0.00006 | 0.00025 | 0.00015 | 0.00011 | 0.00011 | 0.00007 | 0.00005 |
| Costs (per person, USD) |  |  |  |  |  |  |  |  |  |
| Cost of immunization program | 4.4 | 11.3 | -7.0 | 5.8 | 15.2 | -9.4 | 5.8 | 15.2 | -9.4 |
| Productivity lost | 0.5 | 0.3 | 0.2 | 0.8 | 0.5 | 0.3 | - | - | - |
| Cost of treatments | 9.5 | 7.9 | 1.7 | 17.3 | 14.3 | 3.1 | 37.4 | 31.2 | 6.2 |

\*Differences of outcomes between IIV and RIV were calculated as the values of IIV minus the values of RIV. Abbreviations: IIV, inactivated influenza vaccine; RIV, recombinant influenza vaccine; QALYs, quality-adjusted life years; ICER, incremental cost-effectiveness ratio; USD, United States Dollar; ICU, intensive care unit.

Among adults aged ≥65 years, RIV was estimated to prevent 10,157 influenza cases, 1,595 outpatient visits, 776 hospitalizations without ICU admission, 11 hospitalizations with ICU admission, and 17 deaths compared with IIV. Per person, QALY gains were 0.00010 from fewer nonfatal episodes and 0.00011 from deaths averted. Vaccination costs increased by USD9.4 (HKD73.3) per person, while productivity losses and medical costs decreased by USD0.3 (HKD2.3) and USD3.1 (HKD24.2) per person, respectively. Net total costs were USD6.0 (HKD46.8) higher per person with RIV, with an incremental gain of 0.00021 QALYs.

Among adults aged ≥80 years, RIV was estimated to prevent 2,321 influenza cases, 365 outpatient visits, 427 hospitalizations without ICU admission, 2 hospitalizations with ICU admission, and 4 deaths compared with IIV. QALY gains per person were 0.000010 from fewer nonfatal episodes and 0.00005 from deaths averted. RIV increased vaccination costs by USD9.4 (HKD73.3) per person and reduced medical costs by USD6.2 (HKD48.4) per person, yielding a net increase of USD3.2 (HKD25.0) in total costs and an additional 0.00015 QALYs per person.

Cost-effectiveness results are presented in Table 3. Using three times the 2024 Hong Kong GDP per capita as the WPT threshold (USD162,304 [HKD1,265,970] per QALY gained), RIV was cost-effective compared with IIV in adults aged ≥50, ≥65, and ≥80 years, with ICERs of USD40,659 (HKD317,140), USD29,077 (HKD226,801), and USD21,092 (HKD164,518) per QALY gained, respectively. If using a lower threshold of the GDP per capita, we estimated that RIV is cost-effective in approximately 57.8%, 68.7%, and 79.2% of probabilistic sensitivity analysis simulations for individuals aged ≥50 years, ≥65 years, and ≥80 years, respectively.

**Table 3.** Results for cost-effectiveness analysis (per-person costs and effectiveness).

| Age | Strategies | Total costs (USD) | Total QALYs | Incremental costs (USD) | Incremental QALYs | ICER (USD) |
| --- | --- | --- | --- | --- | --- | --- |
| ≥50 years | IIV | 14.4 | 0.00037 | - | - | - |
|  | RIV | 19.5 | 0.00025 | 5.1 | 0.00012 | 40,659 |
| ≥65 years | IIV | 23.9 | 0.00060 | - | - | - |
|  | RIV | 29.9 | 0.00039 | 6.0 | 0.00021 | 29,077 |
| ≥80 years | IIV | 43.2 | 0.00048 | - | - | - |
|  | RIV | 46.4 | 0.00033 | 3.2 | 0.00015 | 21,092 |

Abbreviations: IIV, inactivated influenza vaccine; RIV, recombinant influenza vaccine; QALYs, quality-adjusted life years; ICER, incremental cost-effectiveness ratio; USD, United States Dollar.

### Deterministic sensitivity analysis

Figure 2 presents the ten most influential parameters based on the sensitivity analyses for adults aged ≥50, ≥65, and ≥80 years. Across all age groups, RIV remained cost-effective as each parameter was varied across its plausible range. For adults aged ≥50 and ≥65 years, relative vaccine efficacy against illness in adults aged ≥65 years was the most influential parameter. When this parameter was set to its upper bound, the ICER decreased to USD26,990 (HKD210,522) per QALY gained for adults aged ≥50 years and USD17,428 (HKD135,938) per QALY gained for adults aged ≥65 years. At the lower bound, the ICER increased to USD113,225 (HKD883,155) per QALY gained and USD125,660 (HKD980,148) per QALY gained, respectively.

**Figure 2.**
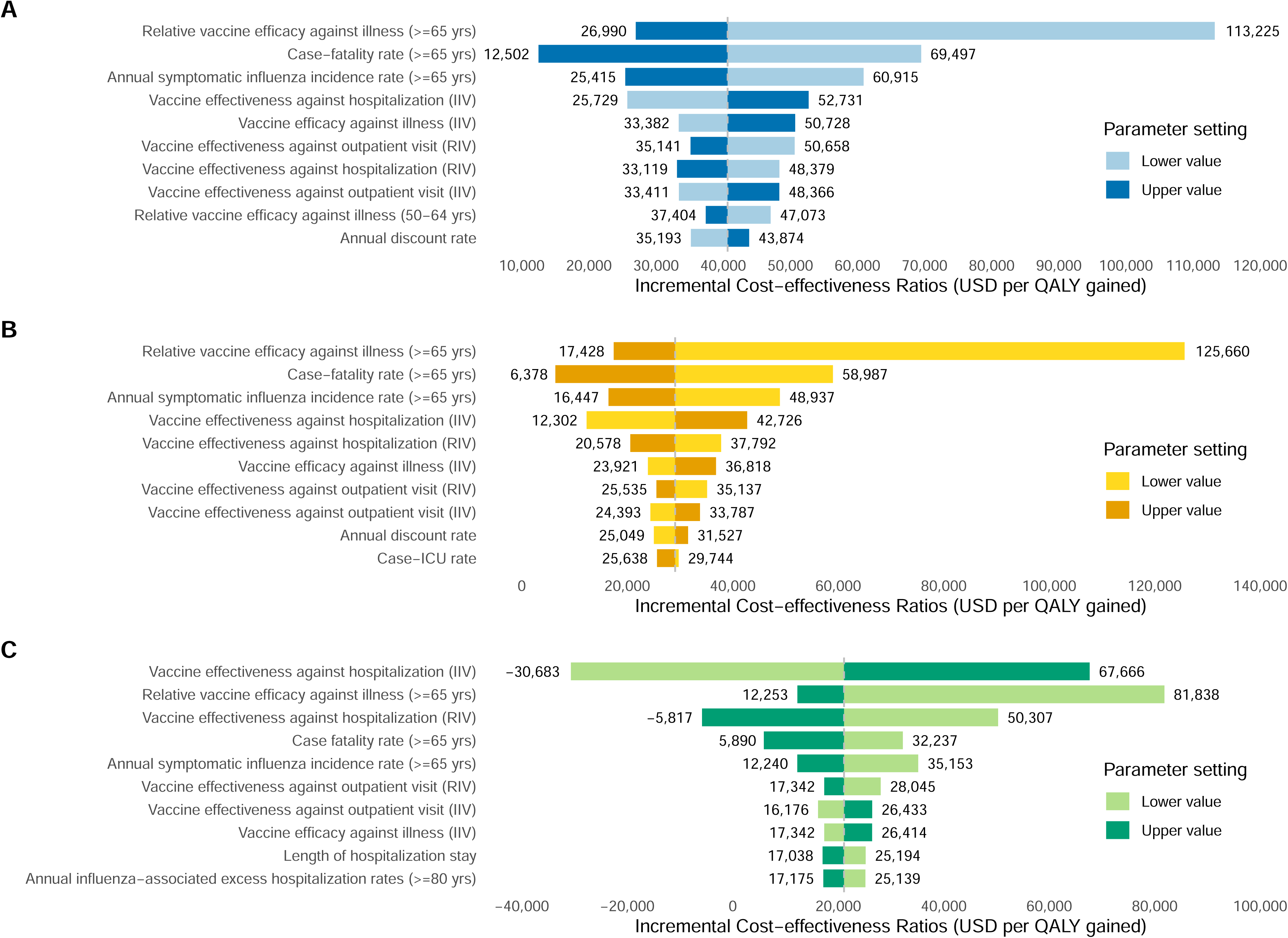
Tornado diagram in the one-way sensitivity analyses of recombinant influenza vaccine (RIV) versus inactivated influenza vaccine (IIV) in adults aged (A) ≥50 years, (B) ≥65 years, (C) ≥80 years. Abbreviations: QALYs: quality-adjusted life-years; ICU, intensive care unit; USD, United States Dollar.

For adults aged ≥80 years, vaccine effectiveness of IIV against hospitalization was the most influential parameter (Figure 2C). When this parameter was at its lower bound, the ICER was USD-30,683 (HKD-239,327) per QALY gained, indicating that total costs of RIV were lower than those of IIV (i.e. RIV was cost𝚦saving). In contrast, the ICER increased to USD67,666 (HKD527,795) per QALY gained when this parameter was at its upper bound.

### Probabilistic sensitivity analysis

The probabilistic sensitivity analysis results are shown in the cost𝚦effectiveness plane (Figure 3) and the cost-effectiveness acceptability curve (Figure 4). At a WTP threshold of three times Hong Kong’s GDP per capita (USD162,304 [HKD1,265,970]), RIV was cost-effective in 97.8%, 97.1%, and 96.7% of 1,000 simulations for adults aged ≥50, ≥65, and ≥80 years, respectively. At two times GDP per capita (USD108,203 [HKD843,980]), these probabilities were 92.1%, 92.6%, and 93.0%. At a threshold of one time GDP per capita (USD54,101 [HKD421,990]), the probabilities were 57.8%, 68.7%, and 79.2%, respectively.

**Figure 3.**
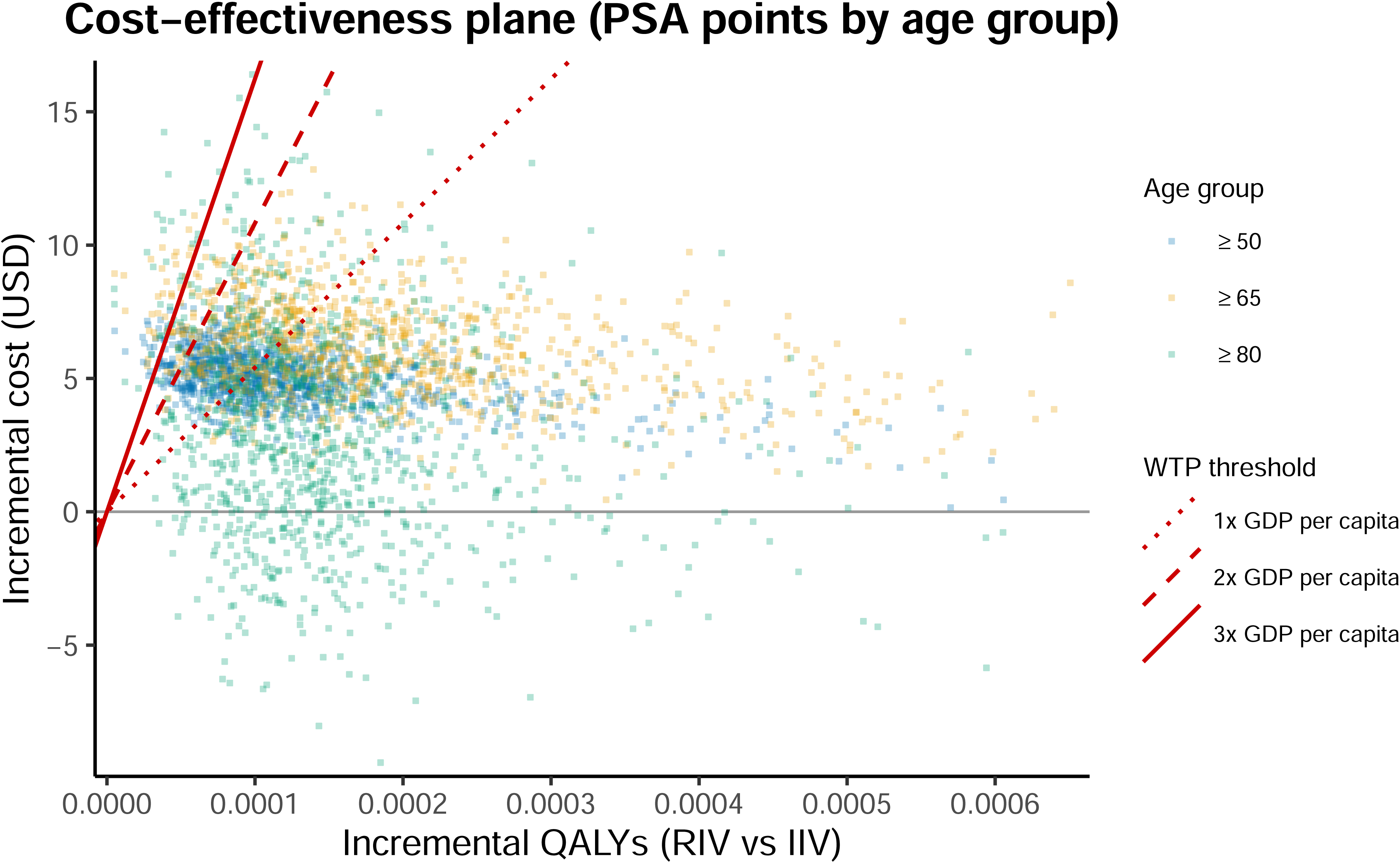
Cost-effectiveness plane displaying 1,000 Monte Carlo simulations of incremental costs and incremental quality-adjusted life years (QALYs) for recombinant influenza vaccine (RIV) versus inactivated influenza vaccine (IIV). The red dotted, dashed, and solid lines denote willingness-to-pay (WTP) thresholds at one, two, and three times Hong Kong’s 2024 gross domestic product (GDP) per capita, respectively: USD54,101 (HKD421,990; 1× GDP); USD108,203 (HKD843,980; 2× GDP); and USD162,304 (HKD1,265,970; 3× GDP).

**Figure 4.**
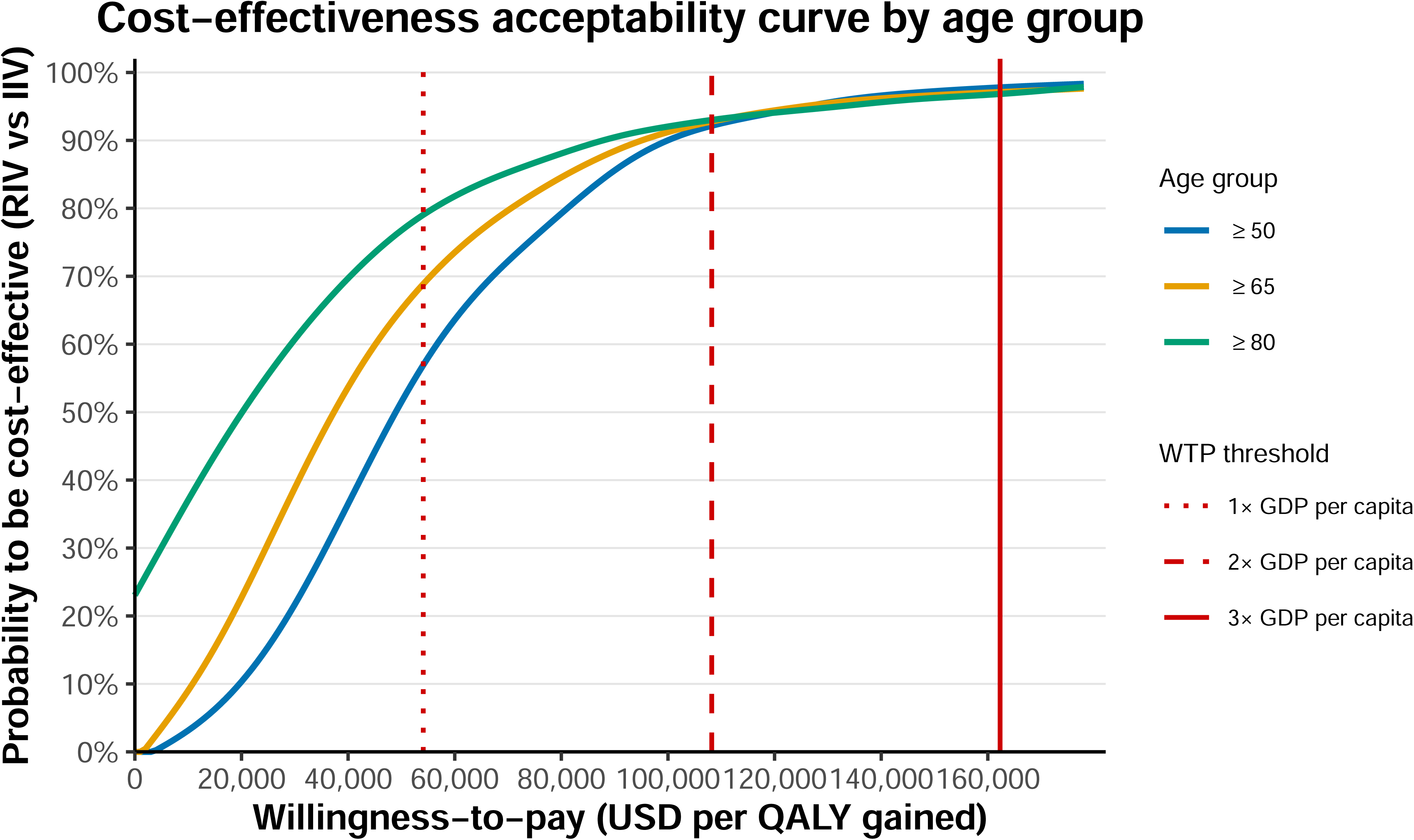
Cost-effectiveness acceptability curve for recombinant influenza vaccine (RIV) compared with inactivated influenza vaccine (IIV). Red lines represent willingness-to-pay thresholds at one (dotted), two (dashed), and three (solid) times Hong Kong’s 2024 gross domestic product (GDP) per capita: USD54,101 (HKD421,990); USD108,203 (HKD843,980); and USD162,304 (HKD1,265,970), respectively.

## Discussion

From a societal perspective, this study used a decision𝚦tree model to evaluate the cost𝚦effectiveness of RIV compared with IIV across age groups in Hong Kong. At a WTP threshold of one to three times the 2024 Hong Kong GDP per capita (USD162,304 [HKD1,265,970]), RIV was estimated to be cost𝚦effective relative to IIV for adults aged ≥50, ≥65, and ≥80 years.

The one-way sensitivity analyses demonstrated that our conclusions are robust to plausible variation in key parameters. Among the parameters tested, the relative vaccine efficacy of RIV versus IIV against symptomatic influenza illness exerted the greatest influence on the ICER. The observed negative association between relative vaccine efficacy and ICER indicates that the cost-effectiveness of RIV improves as its clinical advantage over IIV increases.

The age𝚦stratified analyses provide important guidance for prioritizing RIV use. In the base𝚦case analysis, replacing IIV with RIV in adults aged ≥80 years yielded the lowest ICER, indicating that targeting this oldest age group is the most cost-effective strategy. This aligns with the well-documented increase in influenza-related disease burden with age [1,30,41]. When public health budgets are constrained, prioritizing RIV for the oldest population would maximize health gains per dollar spent. Vaccination with RIV at younger ages (≥50 and ≥65 years) also remained cost-effective, though with less favorable ICERs than in older age strata.

These findings align with and extend current vaccination policy in Hong Kong. RIV is licensed for individuals aged ≥18 years and is preferentially recommended for older adults in residential care homes, although there is no specific publicized subsidized program for Flublok targeting high-risk groups [7]. Prior to this work, there had been no formal economic evaluation of the use of RIV versus IIV in Hong Kong, nor an assessment of how cost𝚦effectiveness varies by age group. By providing age𝚦stratified ICERs based on local epidemiologic and cost data, this study addresses that gap and offers evidence that can inform refinement of existing recommendations, supporting preferential use of RIV in long𝚦term care facilities and its extension to community𝚦dwelling adults from age 50 years when budgets allow.

Our results are consistent with evidence from other high𝚦income settings. A cost𝚦effectiveness analysis conducted in the United States reported an ICER of USD61,329 per QALY gained for RIV versus IIV among adults aged 50–64 years [42]. While higher than our estimates, this finding demonstrates similar cost-effectiveness conclusions across different healthcare systems, influenza epidemiology, and cost structures. This concordance increases confidence that the cost-effectiveness of RIV relative to IIV is not unique to Hong Kong and may be generalizable to other settings with similar demographic and epidemiologic profiles.

This study has several limitations. First, while we prioritized Hong Kong-specific inputs for our model, some parameters were derived from other countries due to limited local evidence. For example, annual influenza𝚦associated excess outpatient rates and age𝚦specific health utility scores were taken from studies in Japan and mainland China. Nonetheless, our sensitivity analyses showed that variation in these parameters had only modest effects on the ICERs, suggesting that our main conclusions are unlikely to be highly sensitive to these parameters. Second, because estimates of RIV vaccine effectiveness against laboratory𝚦confirmed influenza illness are not available, we used vaccine efficacy as a proxy. Vaccine efficacy often differs from vaccine effectiveness observed in real𝚦world settings [32]. When RIV vaccine effectiveness data become available, future analyses should be conducted to provide more precise estimates. Finally, our decision𝚦tree framework does not capture indirect effects such as herd immunity or transmission dynamics. Therefore, the total population𝚦level benefits of RIV may be underestimated.

In conclusion, based on currently available data and from a societal perspective, RIV is cost𝚦effective compared with IIV for adults aged ≥50, ≥65, and ≥80 years in Hong Kong at a WTP threshold of three times GDP per capita. These findings provide local, policy𝚦relevant evidence to support optimization of influenza vaccination strategies in Hong Kong and may inform decision making in other settings with comparable demographic and epidemiologic characteristics.

## Data Availability

All data produced in the present work are contained in the manuscript

## Acknowledgments

The authors thank Julie Au for technical support.

